# Early functional maturation of anti-Spike antibodies predicts SARS-CoV-2 RNA clearance in mild infection

**DOI:** 10.64898/2026.04.30.26352163

**Authors:** Danielle A S Rodrigues, Barbara G de Araujo, Julia Pestana, Lendel Correia da Costa, Matheus Villanueva Andrade, Layla de Freitas, Brenda Nery, Vicente BT Bozza, Luciana Conde, Letícia M Raposo, Andreza Gama, Vinicius Mendes, Elena Cobos, Luiza Mendonça Higa, Rafael M Galliez, Amilcar Tanuri, Orlando C Ferreira, Leda R Castilho, Terezinha M P P Castineiras, Juliana Echevarria, Marcelo T Bozza, André M Vale

## Abstract

SARS-CoV-2 infection exhibits heterogeneous viral clearance kinetics even in individuals with mild disease. The immunological determinants underlying rapid versus prolonged viral RNA detection remain incompletely defined. Here, we performed a three-year longitudinal analysis of 77 healthcare professionals infected with the ancestral Wuhan strain. Participants were stratified according to viral RNA clearance kinetics into non-persistent (≤21 days) and persistent (>21 days) groups. Non-persistent individuals exhibited accelerated seroconversion to Spike and receptor-binding domain (RBD) antigens during the first 11 days after symptom onset. Early antibody responses were characterized by higher functional activity, including significantly greater neutralizing activity against the ancestral strain. In contrast, persistent individuals displayed delayed seroconversion, prolonged IgM responses, and weaker coordination among IgG subclasses, with less synchronized subclass responses, during early infection. Importantly, total immunoglobulin levels did not distinguish the groups. Cross-variant antibody recognition during acute infection was limited and largely strain-focused in both groups. During convalescence, durable anti-Spike IgG responses were maintained independently of persistence status, without significant differences in cross-variant breadth. Vaccination robustly amplified antibody titers, enhanced variant recognition, and sustained high-affinity responses in both groups. Together, our findings demonstrate that the timing and functional quality of early humoral maturation, reflected by neutralizing antibody activity, rather than antibody magnitude or breadth, are key determinants of SARS-CoV-2 RNA clearance in mild infection. These results highlight the importance of early neutralizing antibody generation in shaping acute viral control and the long-term immune architecture.

## Introduction

The coronavirus disease 2019 (COVID-19) emerged in December 2019 as a novel infectious disease caused by severe acute respiratory syndrome coronavirus 2 (SARS-CoV-2) and rapidly evolved into a global pandemic, resulting in millions of deaths worldwide (Lu et al. 2020, WHO 2026). SARS-CoV-2 infection displays marked heterogeneity in both clinical presentation and virological kinetics, ranging from asymptomatic or mild disease to severe respiratory failure (Fajnzylber et al. 2020). In most mild cases, replication-competent virus is no longer detected approximately 9 days after symptom onset; however, viral RNA may remain detectable by RT-qPCR for 14–17 days or longer (Cevik et al. 2021). Prolonged RT-qPCR positivity has been reported even in non-severe infections (Li et al. 2020, Leitao et al. 2021), raising important questions regarding the immunological mechanisms that govern viral RNA persistence and clearance in the respiratory tract.

Although prolonged viral RNA detection in mild disease is not typically associated with worse clinical outcomes, it may reflect differences in host immune dynamics. Early viral control depends on coordinated innate and adaptive immune responses, including timely activation of virus-specific T and B cell compartments (Iwasaki and Medzhitov 2015, Sette and Crotty 2021). In particular, humoral immunity plays a central role in limiting viral spread by neutralizing virions and facilitating Fc-mediated effector functions (Lu et al. 2018). Neutralizing antibodies targeting the Spike (S) glycoprotein, especially the receptor-binding domain (RBD), have been established as key correlates of protection against symptomatic infection and reinfection (Khoury et al. 2021, Gilbert et al. 2022). However, antibody quantity alone does not fully explain variability in infection outcomes, suggesting that qualitative features such as affinity, subclass distribution, and timing of induction may critically influence viral clearance.

In the context of acute SARS-CoV-2 infection, immunological memory in the form of circulating antibodies and memory B cells persists for at least eight months after symptom onset (Dan et al. 2021). Longitudinal studies have demonstrated ongoing antibody evolution and increasing neutralization potency months after primary infection, even in the absence of re-exposure (Gaebler et al. 2021, Wang et al. 2021). These observations underscore the dynamic nature of humoral maturation following SARS-CoV-2 infection and highlight the importance of affinity maturation and clonal evolution in shaping long-term immunity.

Humoral responses to SARS-CoV-2 are predominantly mediated by immunoglobulin G (IgG), particularly the IgG1 subclass and, to a lesser extent, IgG3, with additional contributions from IgA and IgM isotypes (Atyeo et al. 2020, Luo et al. 2021). Early IgM responses typically precede class switching, whereas IgG1 and IgG3 dominate antiviral immunity and are associated with Th1-biased inflammatory environments (Vidarsson et al. 2014). Vaccination studies have shown that the initial IgG response after a single dose is largely characterized by IgG1 and IgG3, while booster immunization increases overall IgG levels and promotes subclass diversification (Farkash et al. 2021, Tejedor Vaquero et al. 2021). Germinal center–dependent maturation processes are central to the generation of high-affinity, class-switched antibodies and durable humoral memory (Victora and Nussenzweig 2012, Crotty 2019), yet the kinetics of these processes during early infection, and their relationship to viral RNA clearance, remain incompletely defined.

The emergence of SARS-CoV-2 variants of concern further complicated the landscape of protective immunity. Primary infection with the ancestral Wuhan strain generally induces strain-focused neutralizing responses, and several variants demonstrate partial escape from antibodies elicited by earlier strains (Greaney et al. 2021, Cele et al. 2022). Repeated antigen exposure through infection or vaccination can enhance neutralization breadth and expand cross-reactive memory B cell clones (Muecksch et al. 2022). However, it remains unclear whether differences in viral persistence during primary infection influence the maturation kinetics or long-term qualitative features of the antibody response.

Despite extensive characterization of humoral responses in SARS-CoV-2 infection, few studies have directly examined how early antibody kinetics and functional quality relate to the duration of viral RNA detection in mild cases. In particular, it remains uncertain whether accelerated generation of high-affinity and neutralizing antibodies contributes to rapid viral clearance, or whether delayed humoral maturation underlies prolonged viral RNA detection.

In our previous longitudinal analysis of a cohort of healthcare professionals from the same population infected during the first epidemic wave, we identified altered innate immune signatures and described differences in anti-Spike antibody binding associated with prolonged viral RNA detection (Montes-Cobos et al. 2023). However, the functional implications of these humoral differences and their relationship to viral clearance kinetics were not fully explored. To address these questions, we performed a comprehensive longitudinal analysis of humoral immune responses in healthcare professionals infected during the first epidemic wave with the ancestral Wuhan strain. By stratifying individuals according to viral RNA clearance kinetics, we investigated how the timing, affinity, subclass distribution, neutralization capacity, and cross-variant recognition of SARS-CoV-2–specific antibodies relate to viral persistence and subsequent immune architecture. Our findings demonstrate that the early generation of high-affinity, neutralizing antibodies is strongly associated with rapid SARS-CoV-2 RNA clearance, whereas delayed humoral maturation characterizes persistent infection without necessarily conferring a measurable advantage in long-term cross-variant immunity.

## Results

### Cohort characterization and definition of viral persistence

The study cohort comprised 77 healthcare professionals (53 women and 24 men; mean age, 42 years) diagnosed with mild to moderate COVID-19, all infected with the ancestral Wuhan strain in March 2020, and followed longitudinally for three years. Participants were recruited at the COVID-19 Screening and Diagnostic Center (CTD-UFRJ) of the Federal University of Rio de Janeiro, where nasopharyngeal swabs and plasma samples were collected for SARS-CoV-2 RNA detection and antibody response measurements, respectively. A total of 369 samples were collected, representing 2–9 time points per participant, spanning from 1 to 1100 days after symptom onset (DASO). Sampling covered phases of active infection (PCR+), convalescence (PCR–), and vaccination (**Fig. 1A-B**). Two control groups were included: a pre-pandemic control group (n=94; samples collected in 2018) and a pandemic negative control group (n=59; RT-qPCR negative and seronegative for Spike- and RBD-specific IgM, IgA, and IgG) (**Fig. 1C**).

**Figure 1:**
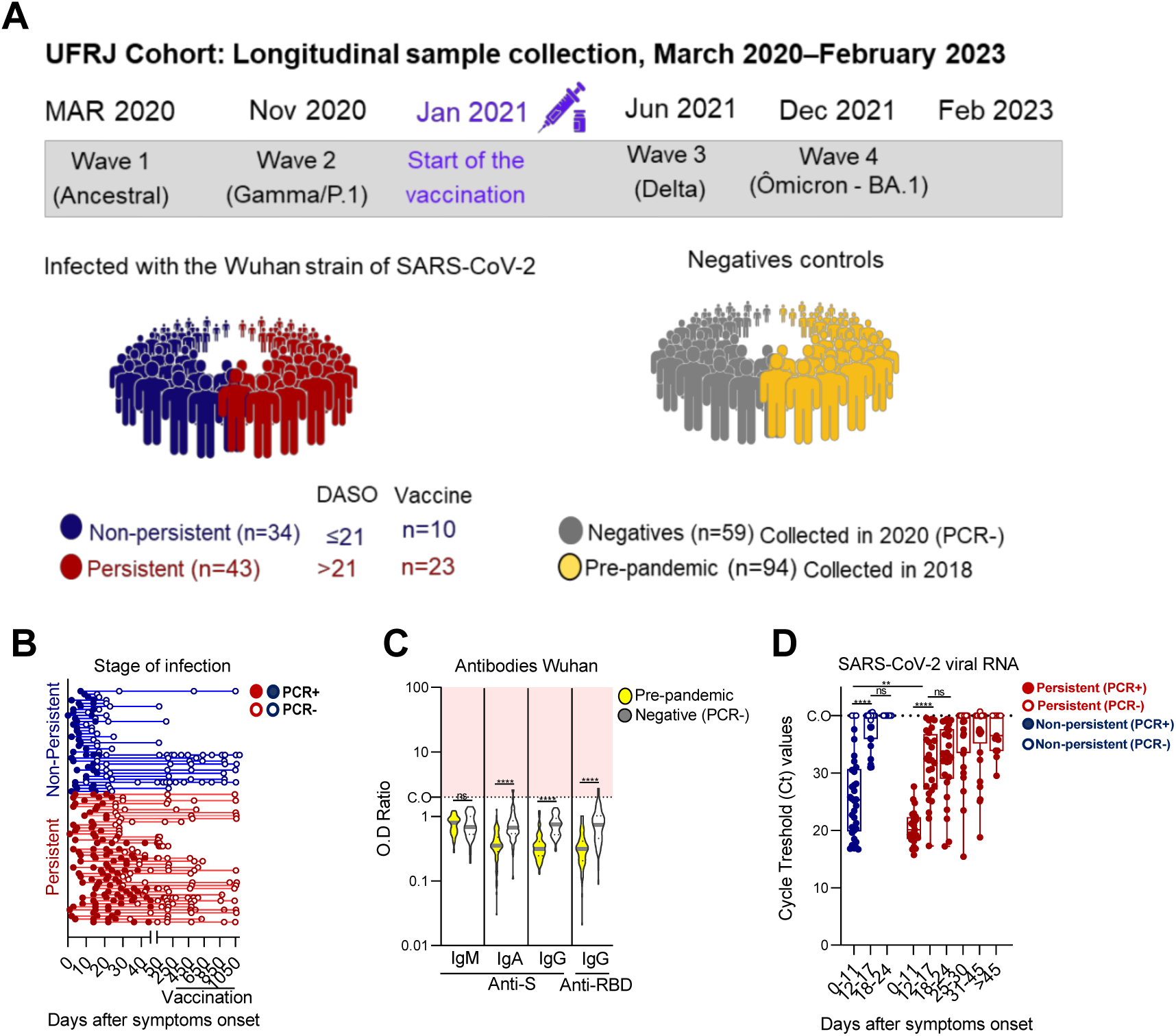
Study design and classification of viral persistence in the UFRJ SARS-CoV-2 cohort. **(A–B)** Timeline and study design of the UFRJ cohort, illustrating the longitudinal collection of blood plasma and nasopharyngeal swab samples from March 2020 to February 2023. The timeline encompasses the predominant SARS-CoV-2 variant waves (Ancestral, Gamma/P.1, Delta, and Omicron BA.1) and the onset of the vaccination campaign in January 2021. Individuals previously infected with the ancestral Wuhan strain were stratified into persistent (n = 43, red) and non-persistent (n = 34, blue) groups based on viral RNA persistence, as defined by DASO, with cutoffs of >21 days and ≤21 days, respectively. Samples were further categorized according to longitudinal PCR status (filled circles=positive; open circles=negative). **(C)** Negative controls comprised PCR-negative individuals sampled in 2020 (n = 59, gray) and pre-pandemic donors sampled in 2018 (n = 94, yellow), all lacking SARS-CoV-2-specific immunoglobulins. **(D)** Cycle threshold (Ct) values for SARS-CoV-2 target genes. Samples with Ct < 40 were considered positive, whereas those with Ct ≥ 40 were considered negative. Statistical significance: ns (not significant); ** (p < 0.01); **** (p < 0.0001).

SARS-CoV-2 infection was monitored through serial nasopharyngeal sampling until viral RNA clearance. Participants were monitored weekly until they achieved negative RT-qPCR results (Ct > 40, indicating no detectable viral RNA). Based on previous CTD-UFRJ data reporting a median viral positivity of 21 DASO (Leitão et al. 2021), this threshold was used to define viral clearance. Individuals were thus classified as *non-persistent* (n = 34; viral clearance ≤21 DASO) or *persistent* (n = 43; remained positive >21 DASO) (**Fig. 1A**, **Table 1**). During the early phase of infection (0–11 DASO), both groups exhibited low Ct values (∼20–30), consistent with high viral loads. Non-persistent individuals displayed viral loads similar to or slightly lower than those of the persistent group. By 21 DASO, all non-persistent participants had cleared the virus (Ct > 40), whereas some persistent individuals remained PCR positive, with Ct values ranging from 25–35 for several weeks and, in some cases, detectable RNA fragments and/or viral particles up to ∼120 DASO (**Fig. 1D**; (Leitao et al. 2021)).

**Table 1:**
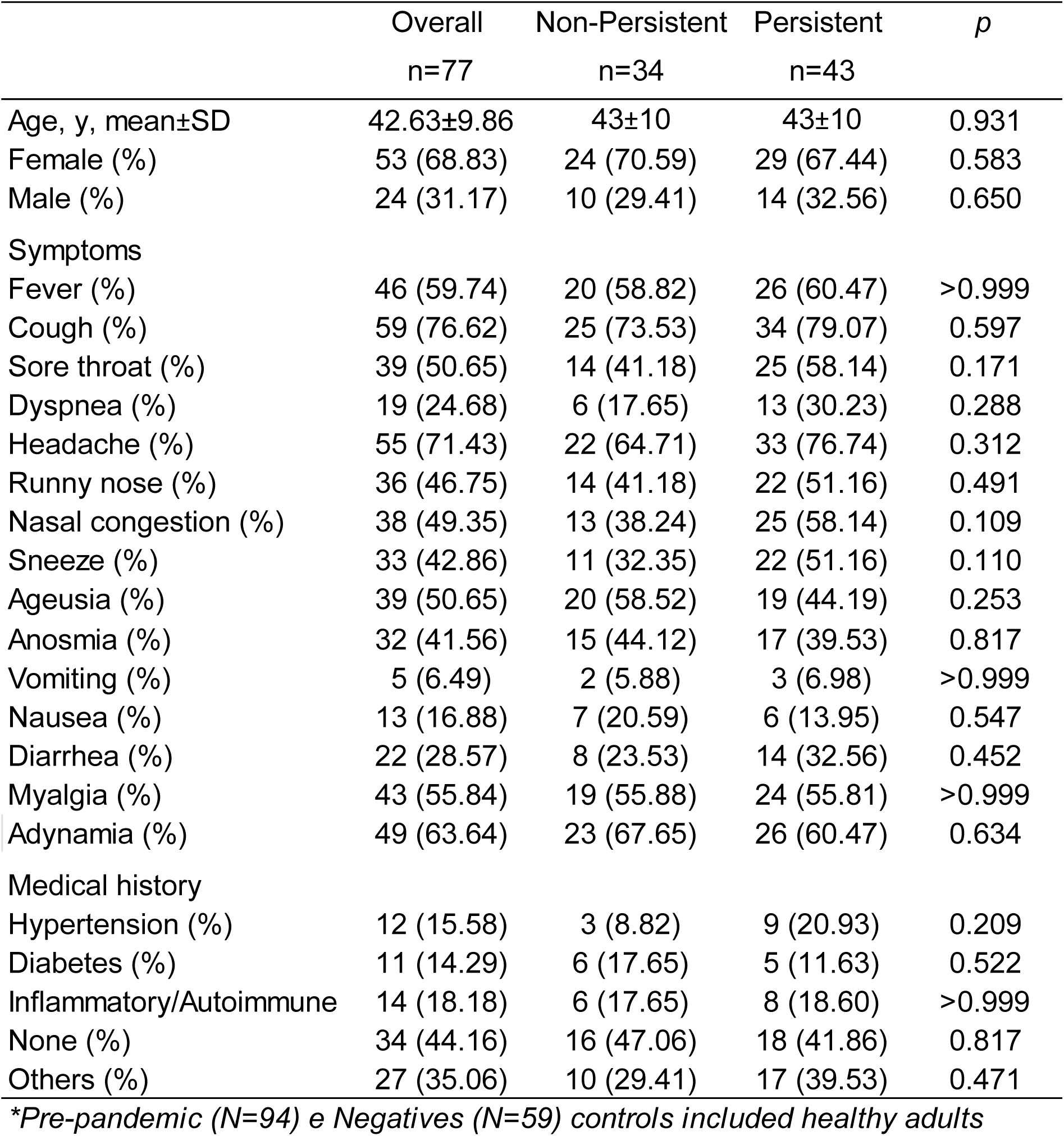
Cohort demographics collected in March 2020.

At disease onset, the most frequent signs and symptoms were cough (76.6%), headache (71.4%), adynamia (63.6%), and fever (59.7%). Sex, age, and comorbidity distributions including hypertension, diabetes, and autoimmune disease were comparable between groups (p > 0.05) (**Table 1**), indicating that prolonged viral RNA detection was not associated with baseline clinical characteristics in this cohort.

### Non-persistent infection is associated with accelerated early seroconversion

To investigate why some individuals rapidly cleared SARS-CoV-2 RNA while others exhibited prolonged RT-qPCR positivity, we analyzed the kinetics of antigen-specific antibody responses during the early phase of infection. We focused particularly on the 0–11 DASO, a critical window for determining subsequent viral clearance dynamics. Although total IgM and IgG levels were comparable between groups (**Suppl. Fig. 1A-B**), marked differences emerged in the timing and magnitude of antigen-specific responses. Most non-persistent individuals mounted detectable and earlier IgM, IgA, and IgG responses against Spike (S), receptor-binding domain (RBD), and nucleocapsid (N) within the first 11 DASO (**Fig. 2A–D**; **Suppl. Fig. 1C**). In contrast, persistent individuals exhibited markedly delayed seroconversion rates, with anti-S and anti-RBD antibodies becoming detectable predominantly between 12–17 DASO, and anti-N responses emerging even later, around 18–24 DASO (**Fig. 2E–H; Suppl. Fig. 1D**). Non-persistent individuals who developed anti-S IgM responses during the early window displayed significantly lower viral loads (i.e., higher CT values), compared to persistent counterparts (**Fig. 2I**). A similar pattern was observed for anti-S, anti-RBD, or anti-N IgG responses, where early IgG positivity correlated with more efficient viral clearance in the non-persistent group (**Fig. 2J–L, Suppl. Fig. 1E**). Together, these results suggest that early seroconversion against key SARS-Cov-2 antigens in non-persistent subjects contributes to improved viral control.

**Figure 2:**
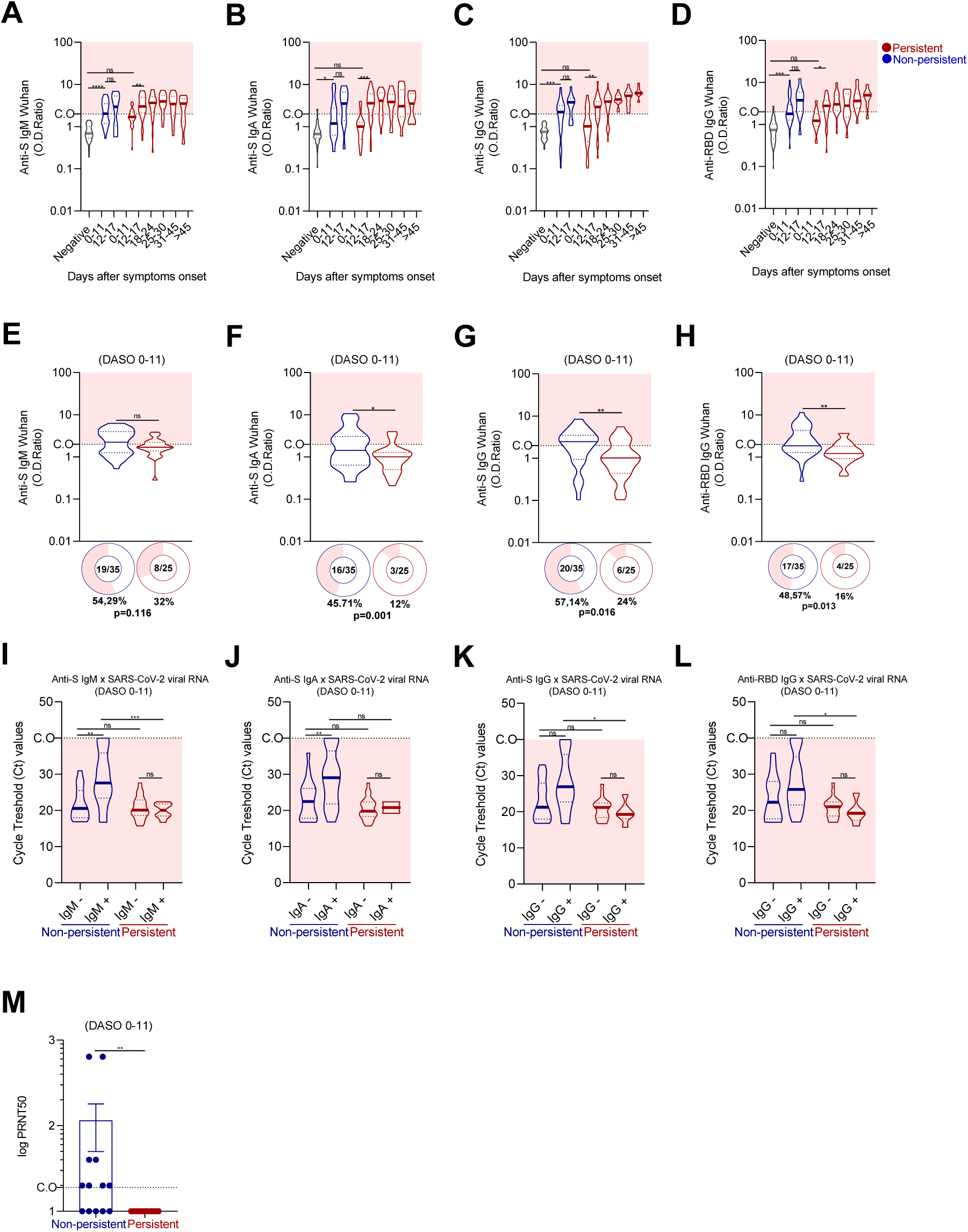
Viral persistence is associated with distinct early antibody responses to SARS-CoV-2. **(A–D)** Distribution of SARS-CoV-2 antigen-specific antibody responses (anti-S IgM, IgA, and IgG; anti-RBD IgG) in individuals previously infected with the ancestral Wuhan strain. Participants were stratified into persistent (red) and non-persistent (blue) groups based on viral RNA persistence, as defined by DASO, during the PCR-positive phase. PCR-negative controls are shown in gray. Relative antibody levels (OD ratio) were calculated by dividing the optical density by the cutoff value (CO). The cutoff was established as the mean absorbance of negative controls plus one standard deviation within each ELISA plate. OD ratios above the cutoff were considered positive, whereas those below were considered negative. **(E–H)** Donut charts showing the proportion and percentage of seropositive versus seronegative individuals in the persistent (red, n = 25) and non-persistent (blue, n = 35) groups during the early infection phase (0–11DASO). **(I–L)** Association between specific antibody seroconversion (anti-S IgM, IgA, IgG, and anti-RBD IgG) and PCR cycle threshold (Ct) values. Samples collected between 0 and 11 DASO were categorized based on the presence (Ig⁺) or absence (Ig⁻) of Spike- or RBD-specific antibodies. **(M)** Serum neutralizing activity measured by PRNT against the ancestral SARS-CoV-2 Wuhan strain. Comparison of PRNT₅₀ titers between non-persistent (blue, n = 13) and persistent (red, n = 13) individuals during the early phase (0–11 DASO). Statistical significance between groups and relative to negative controls: ns (not significant); * (p < 0.05); ** (p < 0.01); *** (p < 0.001); **** (p < 0.0001).

### Early antibody affinity and neutralization capacity predict viral clearance

Given the differences in seroconversion kinetics, we next evaluated whether qualitative features of the antibody response differed between groups during early infection. Using an indirect estimate of antibody affinity based on a multiplex neutralization assay performed in a subset of individuals, non-persistent individuals exhibited higher-affinity anti-Spike antibodies during the early phase compared to persistent individuals **(Suppl. Fig. 1F**). Consistently, sera from non-persistent subjects demonstrated greater neutralizing activity in PRNT assays against the ancestral Wuhan strain (**Fig. 2M**). These findings indicate that not only the timing but also the quality of the early humoral response distinguishes non-persistent from persistent infection. Thus, the early generation of high-affinity neutralizing antibodies likely contributes to rapid viral clearance.

### Humoral coordination and IgG subclass dynamics differ between persistent and non-persistent infection

Protein antigens typically induce T cell–dependent antibody responses dominated by IgG1 and IgG3 subclasses (Vidarsson et al. 2014). Consistent with this pattern, the anti-Spike response in both groups was primarily mediated by IgG1. However, IgG1 appeared earlier in non-persistent individuals (12–17 DASO) and later in persistent individuals (18–24 DASO) (**Fig. 3A**). IgG3 was detected predominantly in persistent individuals after 18–24 DASO, whereas IgG2 and IgG4 remained largely undetectable in both groups across time (**Fig. 3B–D**).

**Figure 3:**
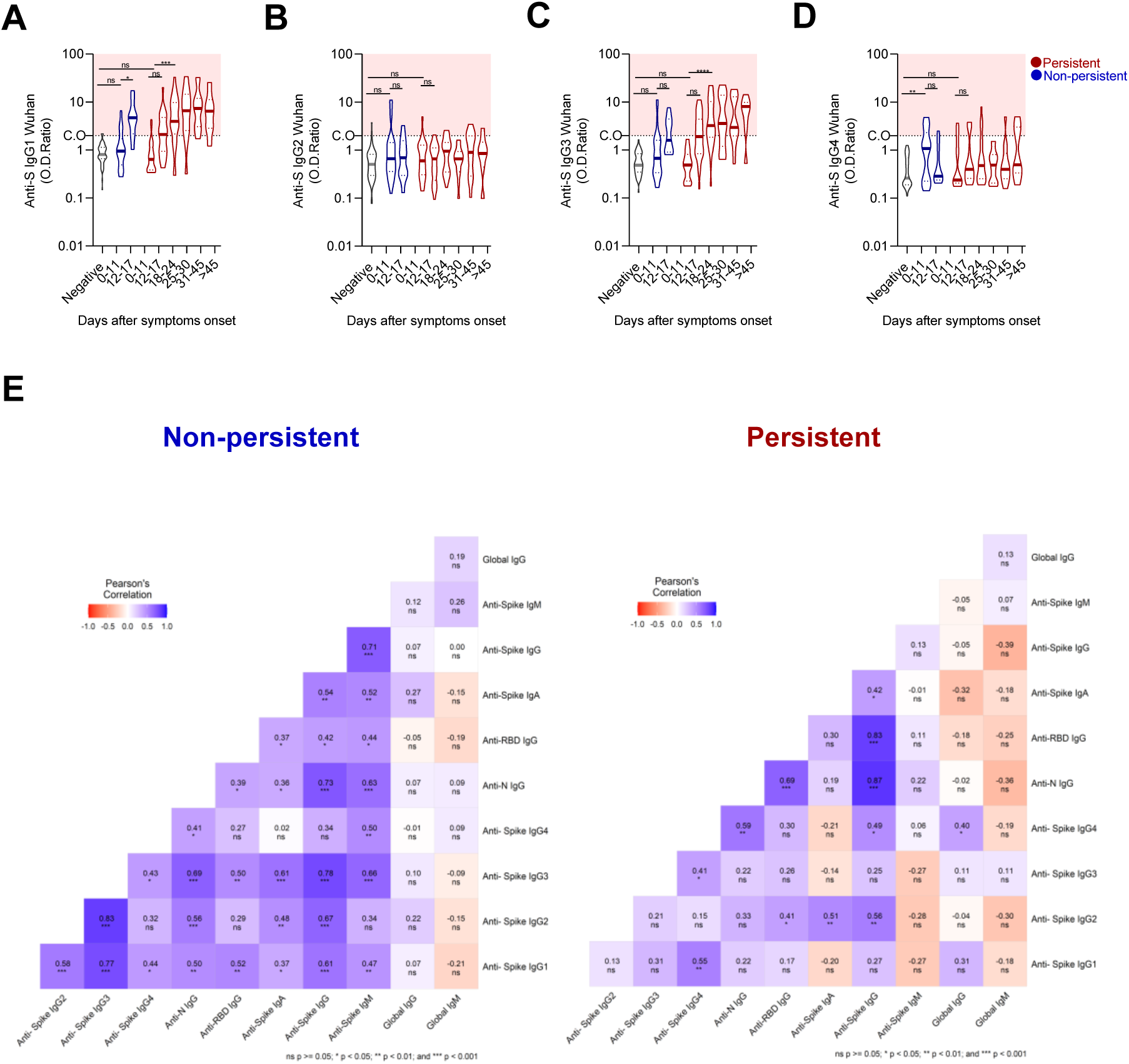
Humoral coordination and IgG subclass dynamics differ between persistent and non-persistent infection. **(A–D)** Longitudinal kinetics of SARS-CoV-2 antigen-specific antibody responses in individuals previously infected with the Wuhan strain. The panels display the levels of anti-Spike IgG subclasses (IgG1–4) measured at various time points after symptom onset. Participants are stratified into non-persistent (blue) and persistent (red) groups based on DASO classification, with PCR-negative controls shown in gray. **(E)** Correlation matrices displaying Pearson correlation coefficients between total immunoglobulins and antigen-specific antibodies (anti-Spike IgG1–4 subclasses, anti-Spike IgA/IgM/IgG, anti-RBD IgG, anti-N IgG, and total IgG/IgM) in non-persistent (left, n = 32) and persistent (right, n = 32) individuals. Color intensity and hue indicate the strength and direction of the correlations (red = negative; blue = positive). Pearson correlation coefficients (r) are shown. Statistical significance: ns (not significant, p ≥ 0.05); * (p < 0.05); ** (p < 0.01); *** (p < 0.001); **** (p < 0.0001).

To assess the coordination of the humoral response, correlation analyses were performed among antigen-specific antibodies and IgG subclasses. Non-persistent individuals exhibited a highly integrated antibody profile, with strong positive correlations among anti-Spike IgG subclasses (IgG2-IgG3, r=0.83; IgG1-IgG3, r=0.77; IgG1-IgG2, r=0.58; all p<0.001), as well as consistent correlations between anti-Spike, anti-RBD, and anti-N IgG responses. In contrast, persistent individuals showed a less coordinated correlation structure, with fewer and weaker associations among IgG subclasses and antigen-specific responses, suggesting reduced synchronization in IgG subclass maturation (**Fig. 3E**). A non-significant trend toward inverse correlations between total IgM and IgG responses was observed in persistent individuals, a pattern not evident in non-persistent subjects (**Fig. 3E**). Although these associations did not reach statistical significance, they are consistent with a less coordinated humoral profile during early infection. These results indicate that coordinated maturation of IgG subclass responses is a feature of effective early humoral immunity underlying viral clearance.

### Primary infection induces durable but subclass-restricted humoral immunity

To determine whether viral persistence influenced long-term immunity, we evaluated antibody responses during the peri-infection phase (first PCR-negative sample) and late convalescence (>70 DASO). Anti-Spike IgM declined rapidly after viral clearance and became undetectable during late convalescence (**Fig. 4A**). Anti-Spike IgA persisted longer but remained less prominent than IgG responses (**Fig. 4B–D**). Both groups maintained robust anti-Spike and anti-RBD IgG responses during late follow-up, indicating durable humoral immunity independent of persistence status (**Fig. 4C–D**).

**Figure 4:**
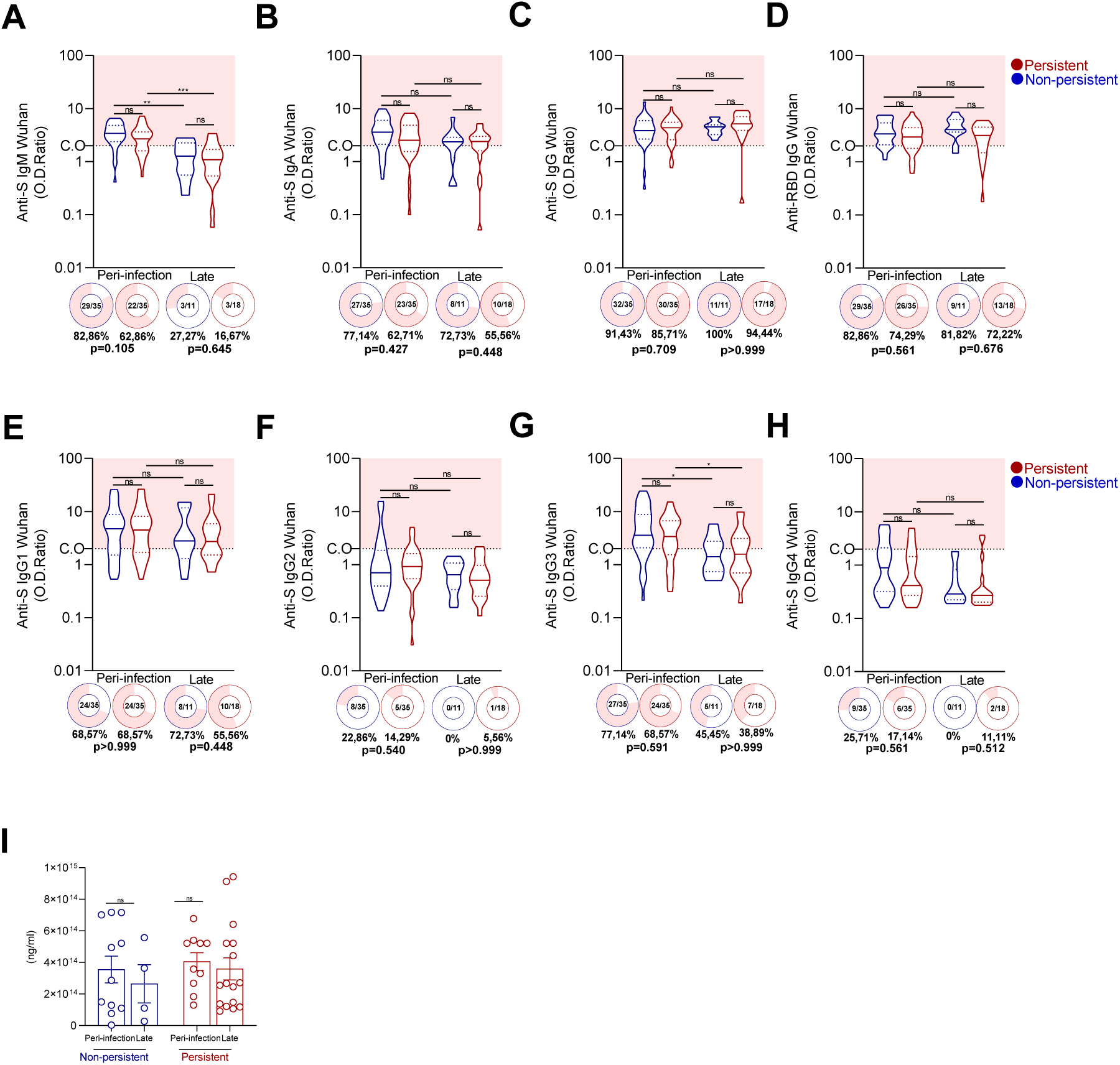
Sustained anti-Spike IgG and high-affinity antibodies despite viral RNA persistence. **(A–D)** Relative levels (OD ratio) of anti-Spike (anti-S) IgM, IgA, IgG, and anti-RBD IgG in non-persistent (blue) and persistent (red) individuals during the peri-infection and late convalescence phases. Relative antibody levels (OD ratio) were calculated by dividing the optical density by the cutoff value (CO). The cutoff was established as the mean absorbance of negative controls plus one standard deviation within each ELISA plate. OD ratios above the cutoff were considered positive, whereas those below were considered negative. Donut charts below each violin plot show the number and percentage of seropositive individuals, with p-values indicating statistical comparisons of seroprevalence between groups. **(E–H)** Distribution and seroconversion rates of anti-S IgG subclasses (IgG1, IgG2, IgG3, and IgG4) across the same clinical groups and time points, evaluated using the same criteria applied to the primary isotypes. **(I)** Concentration (ng/mL) of high-affinity anti-Spike antibodies estimated by a multiplex ACE2-blocking assay in non-persistent and persistent groups during both clinical phases. Statistical significance for comparisons between time points and groups is indicated as follows: ns (not significant); * (p < 0.05); ** (p < 0.01); *** (p < 0.001).

In both groups, IgG1 remained the predominant and most persistent subclass (**Fig. 4E**), whereas IgG2 and IgG4 showed minimal seroconversion (**Fig. 4F, H**). IgG3 anti-S declined over time (**Fig. 4G**). Anti-N IgG levels decreased from the peri-infection to the late phase in the non-persistent individuals (*p* < 0.01), whereas a similar trend did not reach statistical significance in the persistent group. However, inter-group anti-N IgG levels remained comparable at both time points (**Suppl. Fig. 2A**). Importantly, high-affinity anti-Spike antibodies were maintained in both groups during late convalescence (**Fig. 4I**). Individuals with persistent infection exhibited significantly higher total IgM levels during the peri-infection phase compared to non-persistent subjects, consistent with extended antigen exposure during prolonged RNA detection (**Suppl. Fig. 2B**). Nevertheless, total IgG levels remained stable across both phases **(Suppl. Fig. 2C)**. These results suggest that durable humoral immunity is established independently of viral persistence, despite differences in early infection dynamics.

### Early humoral responses are strain-focused and show limited cross-variant recognition

Throughout the COVID-19 pandemic, the emergence of SARS-CoV-2 variants raised the question of whether antibodies elicited by primary infection with the ancestral Wuhan strain could confer cross-reactivity. To address this, we evaluated anti-Spike IgG binding to Spike proteins from major variants of concern (Gamma, Beta, Delta, and Omicron). During the PCR-positive phase, binding to variant Spike proteins was generally low in both persistent and non-persistent individuals (**Fig. 5A–D**). Although isolated statistical differences were observed at specific time points, no consistent pattern of enhanced cross-variant recognition was detected in either group.

**Figure 5:**
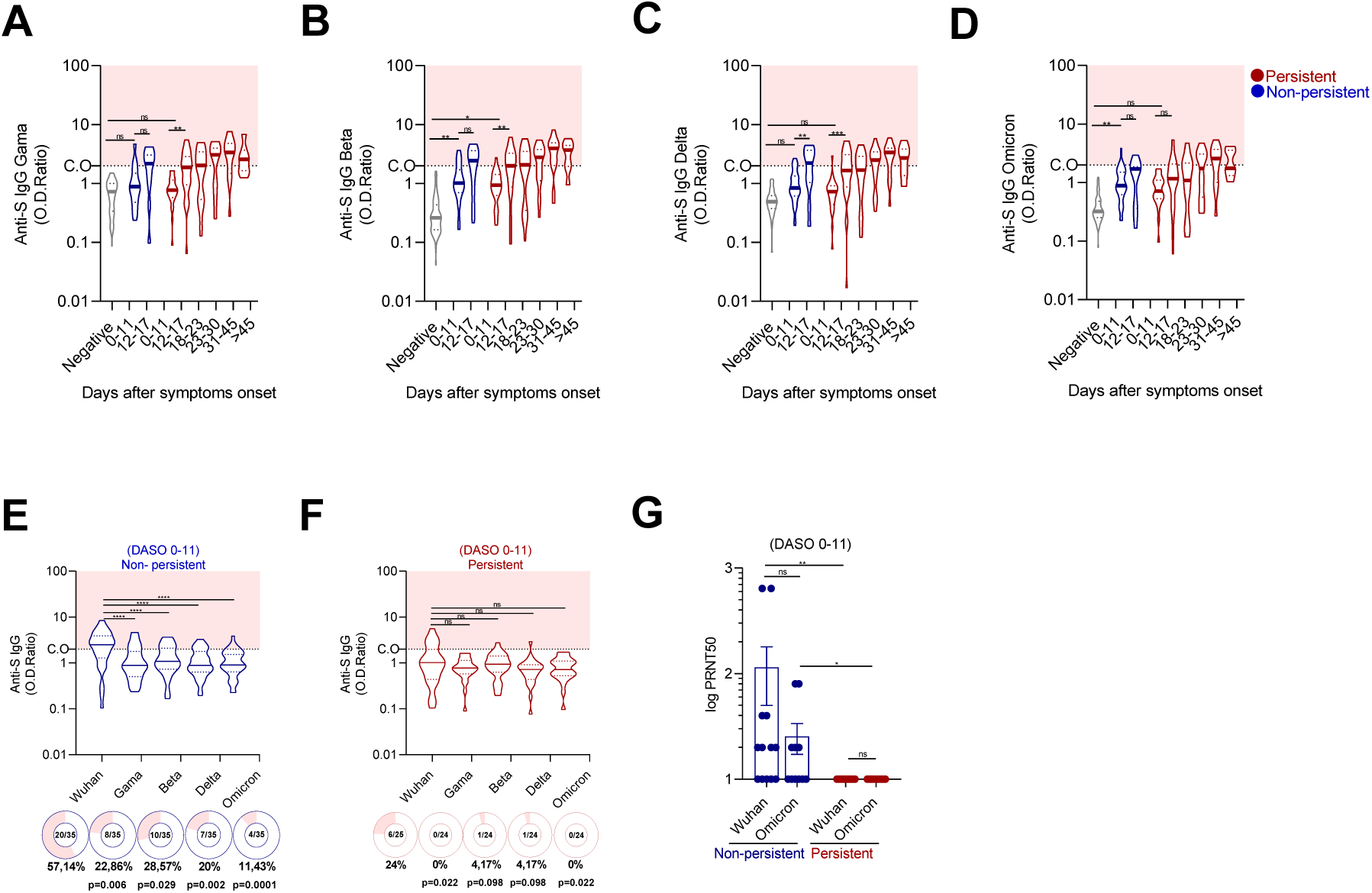
Dynamics of cross-reactive antibodies and early neutralizing capacity against SARS-CoV-2 variants. **(A–D)** Longitudinal kinetics of cross-reactive anti-Spike IgG antibodies against the Gamma, Beta, Delta, and Omicron variants during the PCR-positive phase. Samples are stratified by DASO. PCR-negative controls are shown in gray. Relative antibody levels (OD ratio) were calculated by dividing the optical density by the cutoff value (CO). The cutoff was established as the mean absorbance of negative controls plus one standard deviation within each ELISA plate. OD ratios above the cutoff were considered positive, whereas those below were considered negative. **(E–F)** Early cross-reactive anti-S IgG responses (0–11 DASO) against the ancestral Wuhan strain and variants of concern in non-persistent (n = 35) and persistent (n = 25) groups. Donut charts below the plots show the number and percentage of seropositive individuals for each variant, with p-values indicating statistical comparisons of seroprevalence relative to the ancestral strain. **(G)** Early serum neutralizing antibody activity (0–11 DASO) against the Wuhan strain and the Omicron variant in non-persistent (n = 13) and persistent (n = 12) individuals, expressed as PRNT₅₀. Statistical significance: ns (not significant); * (p < 0.05); ** (p < 0.01); *** (p < 0.001); **** (p < 0.0001).

We next focused on the early phase (0–11 DASO), a critical window for viral clearance. Individuals with non-persistent infection mounted a robust IgG response against the ancestral Spike during this period. However, binding to variant Spike proteins was significantly reduced compared with the ancestral strain, indicating a predominantly strain-specific response (**Fig. 5E**). In contrast, persistent individuals exhibited weaker early recognition of the ancestral antigen and minimal reactivity to variant Spike proteins (**Fig. 5F**). Analysis of anti-RBD IgG responses yielded a similar pattern. In non-persistent individuals, early binding was strongest to the ancestral RBD and declined markedly for Gamma, Delta, and Omicron (**Suppl. Fig. 3A**). Persistent individuals showed low overall reactivity, with limited evidence of variant recognition during this early period (**Suppl. Fig. 3B**). Direct comparisons between groups confirmed that cross-variant IgG binding did not significantly differ between persistent and non-persistent individuals during early infection (**Suppl. Fig. 3C–D**). Consistently, neutralizing activity during this early phase was largely restricted to non-persistent individuals, who exhibited detectable titers predominantly against the ancestral Wuhan strain, with limited cross-neutralization against Omicron (**Fig. 5G**). In contrast, neutralizing activity in persistent individuals was minimal or undetectable. Together, these findings indicate that early viral clearance is associated with rapid generation of functional antibodies directed to the ancestral strain rather than with broad cross-variant humoral recognition.

### Cross-variant seroreactivity during convalescence remains limited and does not significantly differ between groups

We next examined whether cross-variant recognition evolved after viral clearance. During the peri-infection phase (first PCR-negative sample), both persistent and non-persistent individuals displayed strong anti-Spike IgG responses against the ancestral strain (**Fig. 6A–B**). Binding to variant Spikes was detectable in a subset of individuals in both groups. Persistent individuals showed numerically higher proportions of seroreactivity for some variants during this phase. However, direct comparisons between groups did not reveal statistically significant differences (**Fig. 6C**). Analysis of anti-RBD IgG responses during peri-infection phase revealed a more pronounced reduction in variant recognition compared to full Spike, particularly for Omicron (**Suppl. Fig. 4A-B**). Overall, cross-variant recognition remained limited and did not significantly differ between groups (**Suppl. Fig. 4C**).

**Figure 6:**
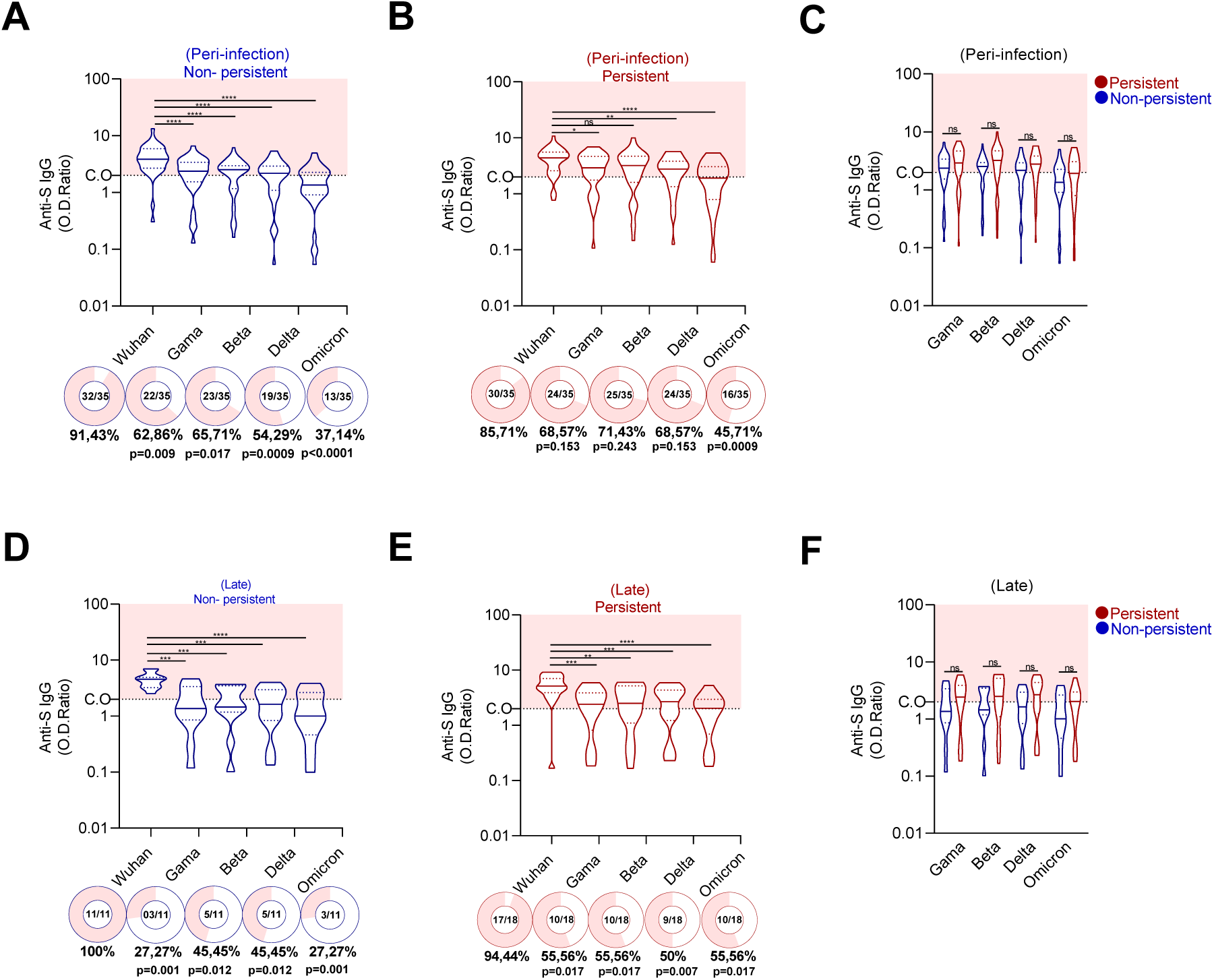
Cross-reactive anti-Spike IgG responses to SARS-CoV-2 variants during peri-infection and late convalescence. **(A–B)** Relative levels (OD ratio) of cross-reactive anti-Spike IgG against the ancestral Wuhan strain and variants (Gamma, Beta, Delta, Omicron) in non-persistent (blue) and persistent (red) individuals during the peri-infection phase. **(C)** Inter-group comparison of antibody levels during the peri-infection phase. **(D–E)** Cross-reactive anti-Spike IgG responses in both groups during the late convalescence phase, analyzed using the same criteria as in (A–B). **(F)** Inter-group comparison during the late phase. Donut charts show the proportion of seropositive individuals, with p-values for group comparisons. Statistical significance: ns; * (p < 0.05); ** (p < 0.01); *** (p < 0.001); **** (p < 0.0001).

During the late convalescence phase, both groups exhibited a reduction in cross-reactive anti-Spike IgG antibodies against variants relative to the ancestral Wuhan strain. However, the persistent group maintained a broader recognition profile, with approximately 50-55% of individuals still recognizing the variants, including the highly divergent Omicron strain (55.56% seropositivity) (**Fig. 6D**). In contrast, the non-persistent group demonstrated a more drastic reduction in cross-reactivity as the genetic distance from the ancestral strain increased, with seropositivity dropping to 27.27% against Omicron (**Fig. 6E**). Notably, despite these distinct intra-group seroprevalence trends, direct quantitative comparisons of cross-reactive antibody levels between persistent and non-persistent individuals revealed no statistically significant differences for any of the tested variants (**Fig. 6F**). Analysis of anti-RBD IgG responses during late follow-up showed similarly reduced cross-variant recognition without significant differences between groups (**Suppl. Fig. 4D–F**).

Collectively, these data demonstrate that primary infection with the ancestral SARS-CoV-2 strain induces durable strain-specific humoral immunity. Cross-variant seroreactivity persists in a subset of individuals but does not significantly differ between persistent and non-persistent groups during follow-up.

### Vaccination sustains and amplifies anti-Spike humoral responses independently of prior viral persistence

We next asked whether prior viral persistence influenced the magnitude or quality of vaccine-induced humoral responses. To address this, we longitudinally analyzed antibody levels before and after vaccination, grouping samples according to days since the start of immunization. Participants received predominantly heterologous vaccine regimens, including CoronaVac, AstraZeneca, Pfizer, and booster doses with mRNA or adenoviral platforms. Following infection resolution, anti-Spike IgM levels declined during convalescence and remained low after vaccination, with occasional increases compatible with possible reinfection events (**Fig. 4A**; **Fig. 7A**). In contrast, vaccination sustained and progressively increased anti-Spike IgA and IgG levels over time in both persistent and non-persistent individuals (**Fig. 7B–C**). Anti-RBD IgG responses followed a similar pattern (**Fig. 7D**). No consistent differences between groups were observed at any evaluated time point.

**Figure 7:**
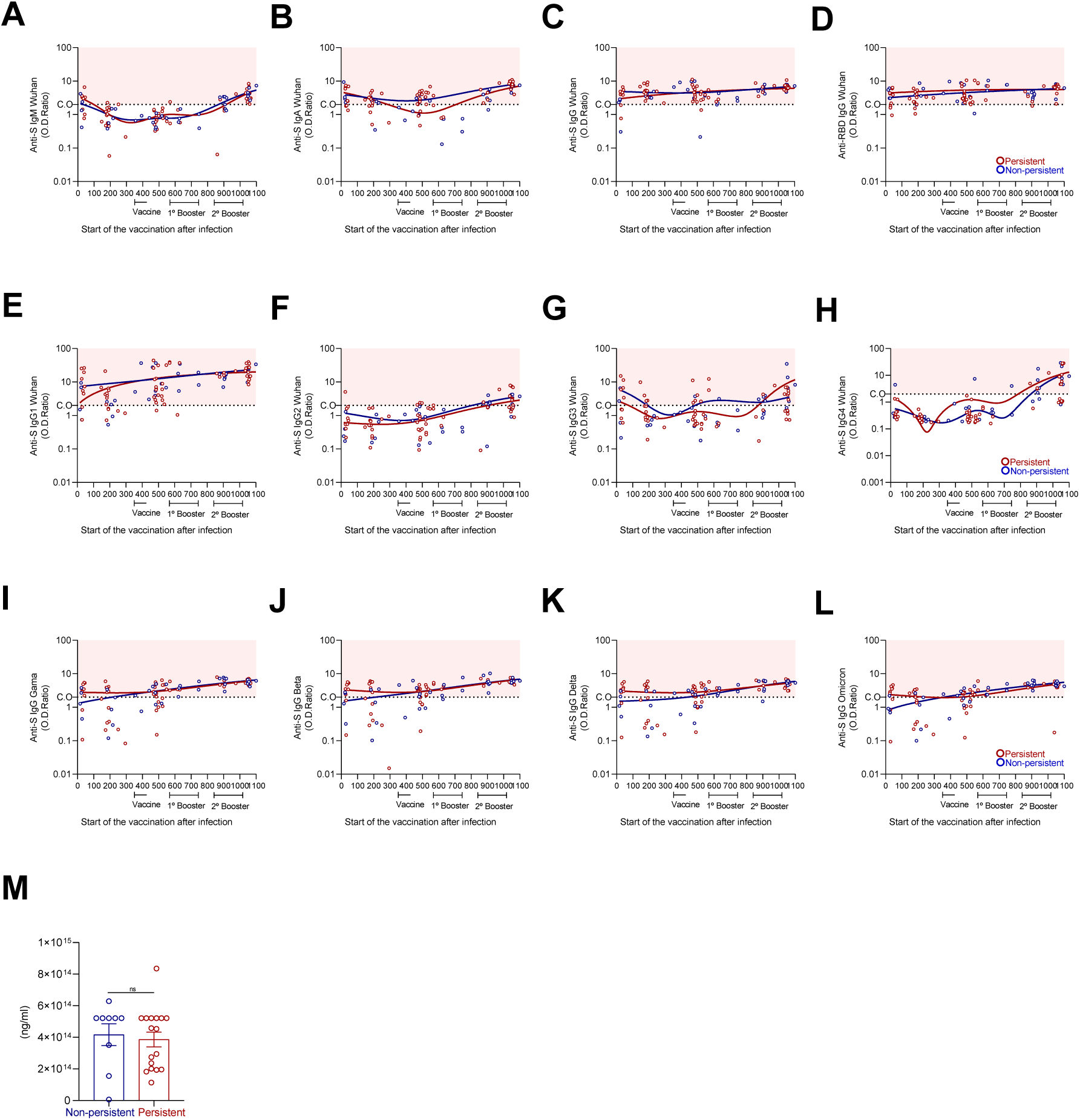
Longitudinal dynamics of SARS-CoV-2-specific antibody responses and affinity following vaccination in convalescent individuals. **(A–D)** Dynamics of anti-Spike IgM, IgA, IgG, and anti-RBD IgG relative levels plotted against days post-vaccination. (E–H) Dynamics of anti-Spike IgG subclasses (IgG1–IgG4) during the post-vaccination period. The plots display the kinetics of antigen-specific antibody levels from the last collection point of the convalescence phase through the post-vaccination period. The study longitudinally followed 23 persistent (red circles) and 10 non-persistent (blue circles) individuals. **(I–L)** Dynamics of anti-Spike IgG against variants of concern (Gamma, Delta, and Omicron) during the post-vaccination period. Relative antibody levels (OD ratio) were calculated by dividing the sample absorbance by the cutoff value (CO). Values below the CO indicate negativity, whereas values above denote seropositivity. The solid kinetic lines represent the expected mean OD ratio derived from a Lowess (locally weighted scatterplot smoothing) regression model. **(M)** Comparison of high-affinity antibody levels estimated by a multiplex ACE2-blocking assay between the non-persistent and persistent groups following vaccination. Statistical significance for all panels: ns (not significant); * (p < 0.05); ** (p < 0.01); *** (p < 0.001); **** (p < 0.0001).

In both groups, analysis of IgG subclasses showed that IgG1 remained the predominant subclass after vaccination (**Fig. 7E**). Modest increases in IgG2, IgG3, and IgG4 were observed following repeated antigen exposure through booster doses (**Fig. 7F–H**). However, subclass kinetics were broadly comparable between groups. Vaccination also enhanced IgG binding to variant Spike proteins in both groups, including Gamma, Beta, Delta, and Omicron (Fig. **7I–L**). Variant recognition increased progressively following primary vaccination and booster doses, with similar trajectories in persistent and non-persistent individuals. Finally, titers of high-affinity anti-Spike antibodies measured after vaccination did not significantly differ between groups (**Fig. 7M**).

Collectively, these data indicate that vaccination robustly amplifies and sustains anti-Spike humoral responses in previously infected individuals, independently of prior viral persistence status.

## Discussion

The determinants of viral clearance during acute SARS-CoV-2 infection remain incompletely defined, particularly in individuals with mild disease. In our previous longitudinal analysis of this cohort, we identified altered innate immune signatures and described differences in anti-Spike binding responses associated with prolonged viral RNA detection (Montes-Cobos et al. 2023). However, the functional implications of these humoral differences and their relationship to viral clearance kinetics remained unresolved. In this longitudinal study spanning three years, we investigated the relationship between humoral immune dynamics and viral persistence in mild-to-moderate COVID-19. Our findings reveal that early coordination and functional quality of the antibody response dictate initial viral clearance. Consistently, viral RNA clearance is associated with the early generation of high-affinity, neutralizing anti-Spike antibodies, whereas total immunoglobulin levels do not distinguish non-persistent from persistent infection. Together, these findings indicate that the timing and functional maturation of the antibody response, rather than its magnitude alone, are key determinants of efficient viral control. Importantly, this early divergence does not compromise the establishment of durable immune memory or the subsequent responsiveness to vaccination in individuals with prolonged viral RNA shedding.

The critical window for establishing viral control occurred within the first 11 days after symptom onset, during which non-persistent individuals mounted accelerated seroconversion to Spike and RBD. This early response was accompanied by higher antibody affinity and greater neutralizing activity directed against the ancestral strain, corroborating previous reports that prompt and robust antibody generation is a hallmark of “quick healers” and early viral controllers (Chen et al. 2020, Long et al. 2020). Early signals of affinity maturation, even when assessed in a subset of individuals, were consistent with functional neutralization, reinforcing the link between antibody quality and viral control.

Building on our previous work (Montes-Cobos et al. 2023), which identified altered innate immune signatures and reported increased antibody binding capacity in individuals with prolonged infection, the present study provides a temporal and functional refinement of these observations. While the previous study primarily focused on early innate responses and antibody features emerging at later time points, it did not resolve the kinetics of early seroconversion. Here, by specifically interrogating the 0–11 DASO window, we demonstrate that accelerated seroconversion and early acquisition of functional antibody activity are key determinants of viral clearance, rather than late increases in antibody binding capacity. Notably, although persistent individuals were previously shown to develop antibodies with enhanced binding capacity at later stages, our data indicate that this delayed maturation does not compensate for the lack of early functional responses required for efficient viral control.

High-affinity antibody production is classically linked to germinal center–dependent maturation (Victora and Nussenzweig 2012, Crotty 2019), and longitudinal analyses of SARS-CoV-2 infection have demonstrated continued antibody evolution over time (Gaebler et al. 2021, Wang et al. 2021). Although we did not directly assess germinal center activity or T follicular helper cell dynamics, the serological patterns observed here are consistent with more rapid functional maturation of the humoral response in individuals who efficiently cleared viral RNA.

In contrast, persistent individuals exhibited delayed seroconversion, prolonged IgM responses, and reduced coordination among IgG subclasses during early infection. These features suggest differences in the timing of class switching and maturation dynamics. Previous longitudinal analyses have demonstrated that severe COVID-19 is characterized by “immunological misfiring,” in which delayed and uncoordinated innate and adaptive immune responses fail to rapidly clear the virus, allowing sustained viral loads that drive clinical deterioration and disease severity (Lucas et al. 2020). Further reinforcing the critical nature of this timeline, the delayed production of neutralizing antibodies, particularly beyond 14 days post-symptom onset, has been directly correlated with impaired viral control and fatal outcomes in cohorts with severe cases (Lucas et al. 2021). Furthermore, severe COVID-19 has been associated with dysregulated extrafollicular activation and disrupted germinal center formation (Kaneko et al. 2020, Woodruff et al. 2020). Although our cohort consisted exclusively of mild-to-moderate cases, the less synchronized humoral profile and delayed kinetics observed in persistent individuals mirror this concept of immune dyscoordination on a milder scale, resulting in prolonged viral RNA shedding rather than severe clinical outcomes. Our findings, therefore, extend the concept that even subtle variations in early humoral maturation kinetics, within clinically mild disease, may influence viral persistence. The association between early neutralizing capacity and clearance supports a model in which prompt functional antibody generation limits ongoing viral replication before prolonged antigenic exposure occurs.

Importantly, viral clearance in this cohort reflects immune responses to the ancestral Wuhan strain, and differences in cross-variant recognition were not associated with viral clearance. During acute infection, antibody responses were largely strain-focused toward the ancestral Wuhan Spike in both groups, and cross-variant neutralization was limited, consistent with previous reports of narrow primary neutralizing repertoires (Greaney et al. 2021, Cele et al. 2022). Clearance was associated with the early emergence of functional antibodies directed against the infecting strain rather than with broader cross-reactivity to divergent variants.

Despite the delayed acute response in the persistent group, our long-term follow-up revealed a convergence in humoral memory. Early in the pandemic, the durability of humoral immunity following mild COVID-19 was a subject of intense debate, with initial studies reporting a rapid decay of circulating anti-SARS-CoV-2 antibodies within the first few months (Ibarrondo et al. 2020, Seow et al. 2020). However, subsequent long-term evaluations demonstrated that after an initial contraction phase, robust neutralizing antibody and IgG responses stabilize and persist in serum and saliva for several months to years (Isho et al. 2020, Iyer et al. 2020, Wajnberg et al. 2020, Dan et al. 2021). In agreement with these latter findings, during convalescence, both persistent and non-persistent individuals maintained durable anti-Spike and anti-RBD IgG responses, predominantly of the IgG1 subclass. IgG1 and IgG3 are hallmarks of Th1-biased antiviral immunity (Vidarsson et al. 2014), and the progressive decline of IgG3 is consistent with its shorter half-life. Persistent individuals exhibited higher total IgM levels during the peri-infection phase, consistent with extended antigen exposure due to prolonged viral RNA shedding. However, this prolonged exposure does not seem to confer a measurable advantage in cross-variant antibody breadth. Although persistent individuals exhibited a numerical trend toward broader cross-variant seroreactivity during later time points, these differences were not statistically significant and did not translate into enhanced functional antibody breadth.

These findings indicate that primary infection with the ancestral strain imprints a predominantly strain-specific humoral memory, and that prolonged viral RNA detection alone is insufficient to drive broad cross-neutralization. While extended antigen exposure may contribute to subtle differences in humoral maturation dynamics, it does not translate into enhanced cross-variant antibody breadth during follow-up. In contrast, repeated antigen exposure through vaccination has been shown to promote the expansion and evolution of cross-reactive memory B cell clones, leading to increased neutralization breadth capable of overcoming major antigenic shifts, such as those presented by the Omicron variant (Cameroni et al. 2022, Muecksch et al. 2022), highlighting that the quality and context of antigen exposure, rather than its duration alone, are critical determinants of humoral diversification.

Vaccination markedly reshaped the humoral landscape in both groups. Immunization with heterologous platforms robustly amplified anti-Spike IgA and IgG levels over time. While IgG1 remained the predominant subclass after vaccination, repeated antigen exposure through booster regimens drove specific subclass dynamics, including modest increases in IgG2, IgG3, and IgG4. Consistent with recent reports demonstrating the emergence of IgG4 alongside sustained overall IgG effector functions following repeated mRNA vaccination (Kaplonek et al. 2026), we observed this specific IgG4 elevation after booster doses. Importantly, the magnitude of this subclass shift is heavily influenced by vaccine platform-specific priming and immune imprinting; repeated mRNA vaccination typically drives a more pronounced IgG4 expansion than other platforms (Kalkeri et al. 2025). This platform dependency likely explains why the IgG4 increases remained modest in our cohort, which received diverse heterologous regimens encompassing inactivated and adenoviral vector vaccines.

Beyond subclass dynamics, booster exposure progressively enhanced IgG binding to variant Spike proteins, including Gamma, Beta, Delta, and Omicron, and maintained high-affinity responses. Crucially, these vaccine-induced amplifications in titers, variant recognition, and antibody affinity occurred with similar trajectories in both persistent and non-persistent individuals. Indeed, it has been clearly demonstrated that while primary SARS-CoV-2 infection in unvaccinated individuals elicits a narrowly focused, strain-specific humoral immunity, it is the integration of vaccination that critically broadens antibody cross-reactivity and enhances Fc effector functions against diverse variants of concern (Richardson et al. 2022). Furthermore, our observation of progressive cross-variant recognition is in agreement with recent findings demonstrating that booster doses are essential for driving affinity maturation and generating robust cross-variant neutralization, particularly against highly divergent lineages like Omicron and its subvariants (Bellusci et al. 2022). These results align with studies showing that vaccination induces robust, cross-reactive IgG responses in previously infected individuals, effectively leveling the immunological playing field regardless of initial infection dynamics (Narowski et al. 2022). Therefore, our data confirm that early differences in infection kinetics and prolonged viral RNA shedding do not constrain subsequent vaccine responsiveness.

This study has limitations. Viral persistence was defined by RT-qPCR detection of viral RNA and does not necessarily reflect replication-competent virus throughout the period of positivity (Cevik et al. 2021). We did not directly evaluate T cell responses, Tfh dynamics, or clonal evolution at the B cell receptor level, which would provide mechanistic confirmation of the maturation differences inferred from serological analyses. Furthermore, all participants were infected with the ancestral strain during the first epidemic wave, which corresponds to the phase in which viral persistence was defined; therefore, immune kinetics may differ in infections caused by later variants with enhanced immune-evasion properties. In addition, information on potential reinfections during the latter follow-up period was not systematically available, which limits our ability to assess their contribution to the evolution of cross-reactive humoral responses over time. The longitudinal design of the cohort inherently led to some loss to follow-up over the three-year period. The finite volume of serum collected at specific time points restricted our capacity to perform all downstream immunological assays (such as neutralization assays and affinity) across the entire cohort. Consequently, the sample size (n) varies across different analyses. Nevertheless, despite this sample attrition, the remaining sample sizes provided sufficient statistical power to identify robust and consistent immunological signatures distinguishing the persistent and non-persistent phenotypes.

In summary, our data support a model in which the timing and functional quality of the early humoral response are decisive for SARS-CoV-2 RNA clearance, even in clinically mild infection. Rapid maturation and early neutralizing activity, rather than antibody magnitude or breadth, are associated with efficient viral control. Viral persistence reflects delayed humoral kinetics but does not necessarily translate into superior long-term variant breadth. Importantly, vaccination consistently elicited stronger and broader humoral responses in both groups, underscoring its critical role in shaping effective and cross-reactive immunity regardless of prior viral persistence status. Together, these findings emphasize that the tempo of antibody functional maturation is a critical parameter shaping both acute infection outcomes and the subsequent immune landscape, linking early humoral kinetics to long-term immune architecture.

## Materials and Methods

### Ethics Statement and Study Cohort

This study was approved by the National Research Ethics Commission (CONEP, Brazil, CAAE: 3016620.0.0000.5257 and number: 3,953,368), and with the consent of all volunteers.

This longitudinal cohort study enrolled 77 healthcare professionals (53 women and 24 men; mean age, 42 years) diagnosed with mild to moderate COVID-19. Blood plasma and nasopharyngeal swab samples were collected between March 2020 and February 2023, encompassing the Ancestral, Gamma, Delta, and Omicron BA.1 waves, as well as the vaccination rollout (started in January 2021). All participants were infected with the ancestral Wuhan strain in March 2020. Recruitment and follow-up were conducted at the COVID-19 Screening and Diagnostic Center (CTD-UFRJ) of the Federal University of Rio de Janeiro. Clinical and demographic data, including symptoms at disease onset (e.g., cough, headache, adynamia, and fever) and comorbidities (e.g., hypertension, diabetes, and autoimmune diseases), were recorded at baseline. Negative controls comprised PCR-negative individuals sampled in 2020 (*n* = 59) and pre-pandemic donors sampled in 2018 (*n* = 94), collected at the State Hematology Institute Hemorio followed a protocol approved 727 by the local ethics committee (CEP Hemorio; approval #4008095).

### Viral RNA Detection and Persistence Stratification

Total viral RNA was isolated from nasopharyngeal swabs using the Maxwell 16 Viral Total Nucleic Acid Purification Kit (Promega) according to the manufacturer’s protocol. SARS-CoV-2 detection was performed using the CDC 2019-nCoV RT-qPCR assay (Integrated DNA Technologies), targeting the N1 and N2 regions of the nucleocapsid gene, with human RNase P serving as an internal control. Reactions were run in parallel on a 7500 Real-Time PCR System (Applied Biosystems). Samples were considered positive when both viral targets amplified with CT ≤ 40 and negative when neither target amplified or amplification occurred with CT > 40. Participants were stratified based on the duration after symptom onset (DASO) into two groups: persistent (*n* = 43; PCR-positive for > 21 days) and non-persistent (*n* = 34; PCR-positive for ≤ 21 days). The first PCR-negative sample collected after viral clearance was defined as the peri-infection phase. The late phase was defined as the period between >70 days after symptom onset (DASO) after viral clearance. This late phase was specifically selected to evaluate the establishment and durability of long-term humoral immune memory prior to the administration of any SARS-CoV-2 vaccines.

### S-UFRJ ELISA for antibodies detection

Antigen-specific antibody responses (IgM, IgA, IgG, and IgG1–4 subclasses) against SARS-CoV-2 Spike (S), Receptor-Binding Domain (RBD), and Nucleocapsid (N) proteins were measured by S-UFRJ ELISA as described previously (Alvim et al. 2022). High-binding 96-well ELISA microplates (Corning) were coated with 50 µL per well of coating solution and incubated overnight at room temperature (RT). The coating solution contained SARS-CoV-2 Spike (S), Receptor-Binding Domain (RBD), or Nucleocapsid (N) proteins diluted in PBS (Gibco) at a final concentration of 4μg/mL. The antigens available for SARS-CoV-2 were: Spike protein SLECC1105 provided by Prof. Leda Castilho (COOPE), the N45Kda by Prof. Ronaldo Mohana (Biophysics) and the RBD by Dr. John Pak from Chan Zuckerberg (BioHub) and Sino Biological Inc. After coating, the wells were washed and blocked with 150 µL of PBS containing 1% BSA (blocking solution) for 1h at RT. Following blocking, 50 µL of serum or plasma samples diluted 1:40 in PBS-1% BSA were added to each well and incubated for 2 h at RT. Plates were washed five times with 150 µL of PBS. Next, 50 µL of HRP-conjugated goat anti-human secondary antibody specific for IgG (Fc), or for IgA, IgM, and IgG subclasses 1–4 (Southern Biotech), diluted 1:8000 in PBS-1% BSA, was added and incubated for 2 h at RT. Plates were again washed five times with 150 µL of PBS. The reaction was developed with 50 µL of TMB substrate (3,3’,5,5’-tetramethylbenzidine; Scienco, Brazil), and stopped by adding 50 µL of 1N HCl. Optical density (O.D.) was measured at 450 nm with background correction at 655 nm using a microplate reader (Bio-Rad). Results are expressed as the ratio between the sample O.D. and the cut-off value. The cut-off was defined as the mean O.D. of the negative controls on the same plate plus one standard deviation, determined from negative control samples tested under identical conditions. Samples with absorbance values below the C.O were classified as negative, while those with values above the C.O were considered positive.

### Plaque reduction neutralization test (PRNT)

Plaque Reduction Neutralization Test (PRNT) was performed for the evaluation of neutralizing antibodies against SARS-CoV-2. Serum samples were heat-inactivated at 56 °C for 30 min, followed by two-fold serial dilutions. The diluted sera were incubated with 100 plaque-forming units (PFU) of SARS-CoV-2 lineage B isolate (SP1, GenBank accession number MT126808.1) or SARS-CoV-2 Omicron lineage BA.1 isolate (EPI_ISL_7699344), for 1 h at 37°C. In addition, virus-only and cell-only controls were included to determine virus infectivity and to monitor cell monolayer integrity, respectively. The virus-serum mixtures were inoculated onto confluent VERO-hACE2 cells seeded in 24-well plates and allowed to adsorb for 1 h at 37°C. After adsorption, the inoculum was removed and replaced with an overlay medium (1.25% carboxymethylcellulose in alpha-MEM supplemented with 1% fetal bovine serum). Cells were further incubated for 3 days, after which the cells were fixed with 4% formaldehyde and stained with 0.25% crystal violet in 20% ethanol for plaque visualization. The plaque reduction neutralization titer was defined as the highest serum dilution resulting in a 50% reduction in plaque counts (PRNT50) when compared to the mean plaque counts of virus-only controls. A PRNT50 titer of ≥ 20 was considered positive for the presence of neutralizing antibodies (Drews et al. 2021, Mahmoud et al. 2021). All experiments involving replication-competent SARS-CoV-2 were conducted in a Biosafety Level 3 (BSL-3) laboratory located in the Laboratório de Virologia Molecular, Universidade Federal do Rio de Janeiro (UFRJ, Brazil).

### Multiplex Surrogate Neutralization Assay (ACE2-blocking)

To evaluate the functional capacity of antibodies to inhibit the Spike-ACE2 interaction, sera were analyzed using the Bio-Plex Pro Human SARS-CoV-2 Neutralization Antibody Assay (BIO-RAD), according to the manufacturer’s instructions. This competitive multiplex assay measures neutralizing antibodies that block the binding of the viral antigens to the human ACE2 receptor.

### Statistical analysis

Statistical analyses were conducted using GraphPad Prism (version 8.0) and R-Studio software. Quantitative continuous variables (such as relative antibody levels) were compared between two independent groups using the Mann–Whitney U test. For multiple comparisons, the Kruskal-Wallis test followed by Dunn’s post-test, or alternatively, a one-way analysis of variance (ANOVA) followed by Tukey’s post-test, was employed depending on data distribution. Differences in categorical variables, such as seropositivity rates, were assessed using Fisher’s exact test. Correlation analyses among antigen-specific antibodies and IgG subclasses were performed using Pearson’s correlation coefficients, as appropriate. Kinetic curves for longitudinal data were fitted using a locally weighted scatterplot smoothing (Lowess) regression model. The number of experimental subjects per group (*n*), the specific statistical tests applied, and summary statistics are detailed in the respective figure legends.

## Data Availability

All data are available in the main text or the supplementary materials.

## Acknowledgments

The authors would like to thank all the study participants for their time and willingness to contribute to this research. Furthermore, we are grateful to Ronaldo Rocha (LAGIIVIR, IMPG-UFRJ) and John Pak (Chan Zuckerberg Biohub) for their expert technical support.

## Author contributions

Conceptualization: DASR, MTB and AMV

Methodology: DASR, LMH, JE and AMV

Investigation: DASR, BGA, JP, LCC, MVA, LF, BN, VTB, EC, LMH, JE, MTB, AT, OCFJr, RMG, TMPPC and AMV

Patient enrollment: RMG and TMPPC

Sample processing and storage: DASR, BGA, LF, LC, AG, VM, EC and JE

Spike proteins production: LRC

Neutralization assays: LCC, MVA and LMH

Intellectual contribution: DASR, MTB and AMV

Analyzed data: DASR, LMR, LCC, MVA, LMH and AMV

Funding acquisition: DASR, MTB and AMV

Resources: DASR, MTB and AMV

Writing—original draft: DASR and AMV

Writing—review & editing: DASR, LC, MTB and AMV

## Data and materials availability

All data are available in the main text or the supplementary materials.

## Supplementary materials

**Supplementary Figure 1:**
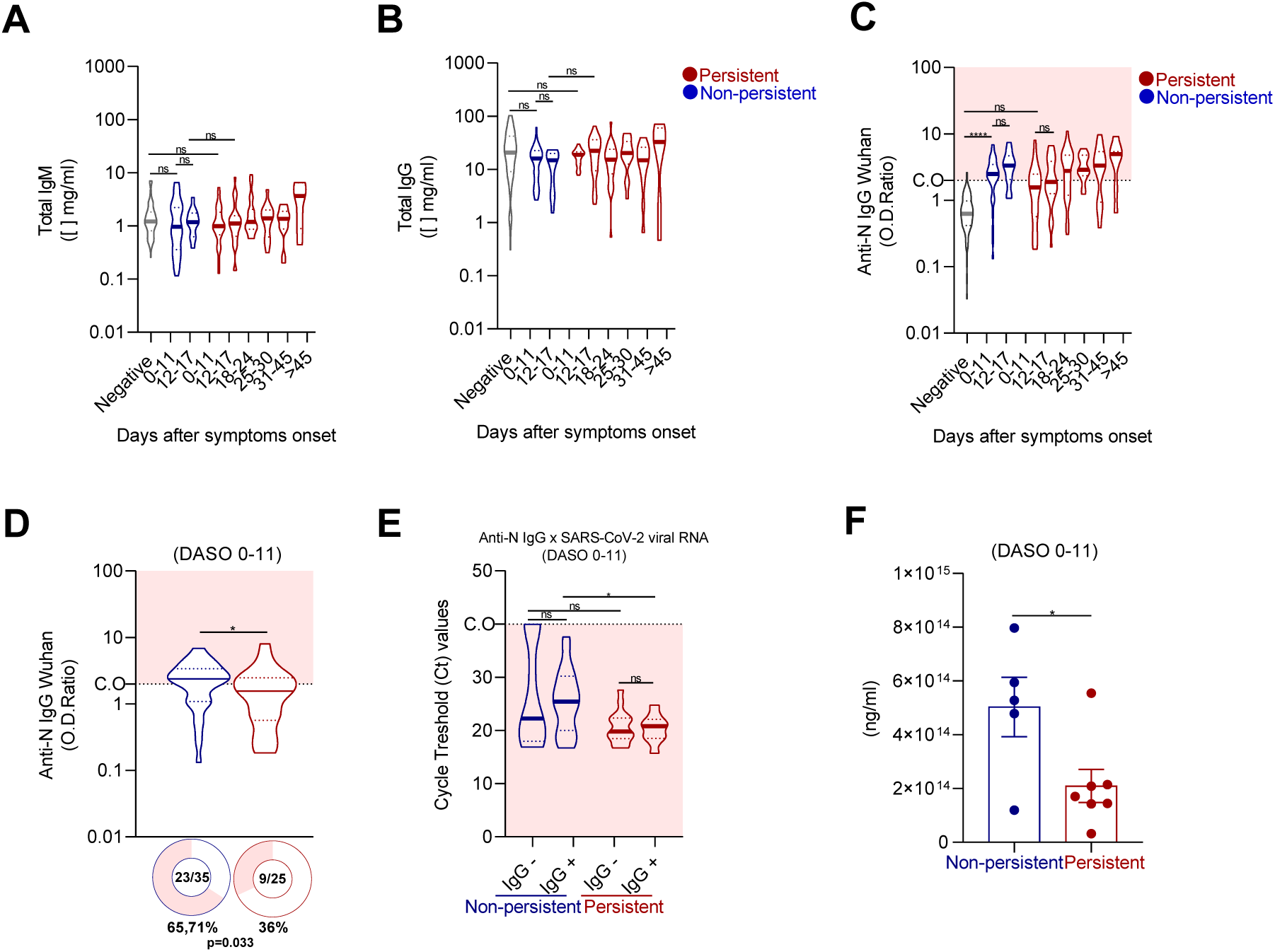
Early humoral responses and antibody affinity in persistent and non-persistent SARS-CoV-2 infection. **(A–B)** Serum concentrations of total IgM and IgG antibodies in individuals infected with SARS-CoV-2. **(C)** Distribution of antigen-specific anti-N IgG antibody responses in individuals previously infected with the ancestral Wuhan strain. Participants were stratified into persistent (red) and non-persistent (blue) groups based on viral RNA persistence, as defined by DASO, during the PCR-positive phase. PCR-negative controls are shown in gray. Relative antibody levels (OD ratio) were calculated by dividing the optical density by the cutoff value (CO). The cutoff was established as the mean absorbance of negative controls plus one standard deviation within each ELISA plate. OD ratios above the cutoff were considered positive, whereas those below were considered negative. **(D)** Donut charts show the proportion of seropositive and seronegative individuals in each group during the early infection phase (0–11 DASO). **(E)** Association between anti-N IgG seroconversion and viral load, represented by PCR cycle threshold (Ct) values. Samples collected between 0 and 11 DASO were categorized by the presence (IgG⁺) or absence (IgG⁻) of N-specific antibodies. **(F)** Comparison of high-affinity anti-Spike antibody titers estimated by a multiplex ACE2-blocking assay in a subset of individuals: persistent (n = 7) and non-persistent (n = 5), during early infection (0–11 DASO). Statistical significance: ns (not significant); * (p < 0.05); **** (p < 0.0001).

**Supplementary Figure 2:**
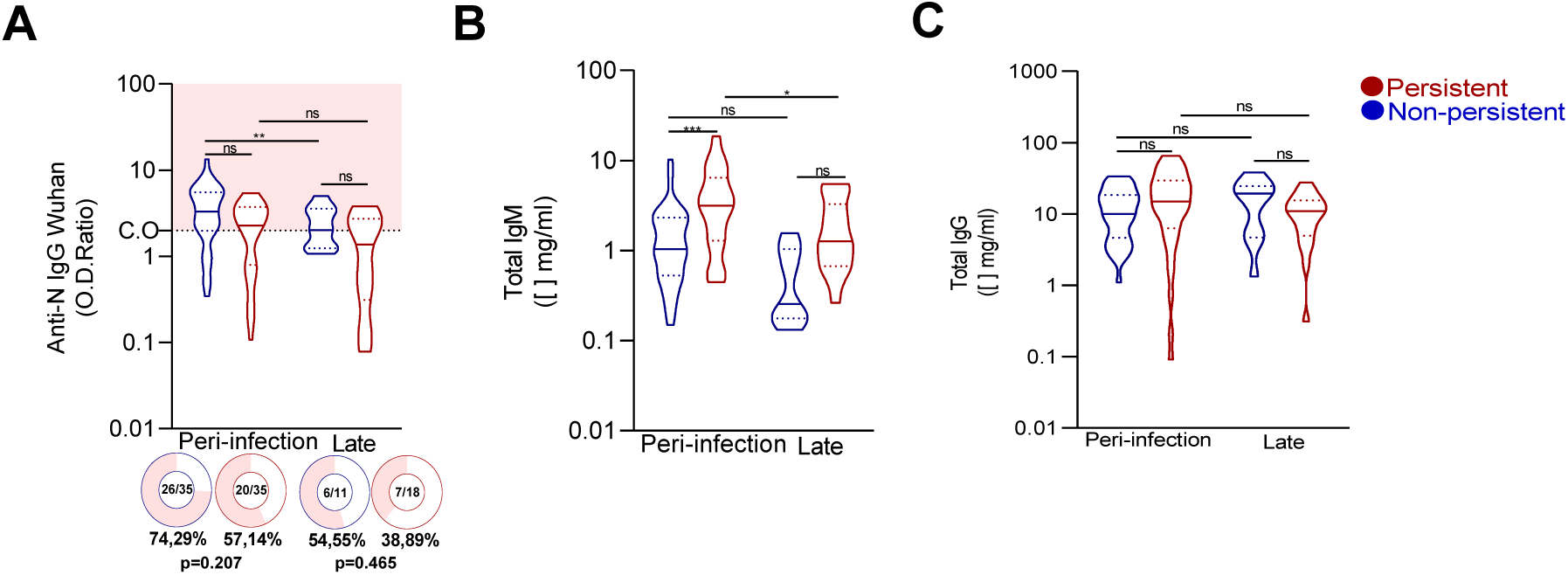
Longitudinal dynamics of anti-Nucleocapsid IgG and total immunoglobulins during peri-infection and late convalescence. **(A)** Relative levels (OD ratio) of anti-Nucleocapsid (anti-N) IgG in non-persistent (blue) and persistent (red) individuals during the peri-infection and late convalescence phases. Donut charts show the proportion of IgG-seropositive individuals in each group, with corresponding p-values indicating statistical comparisons between groups. **(B–C)** Serum concentrations (mg/mL) of total IgM (B) and total IgG (C) in the same clinical groups during the peri-infection and late phases. Statistical significance for comparisons between time points and groups is indicated as follows: ns (not significant); * (p < 0.05); ** (p < 0.01); *** (p < 0.001).

**Supplementary Figure 3:**
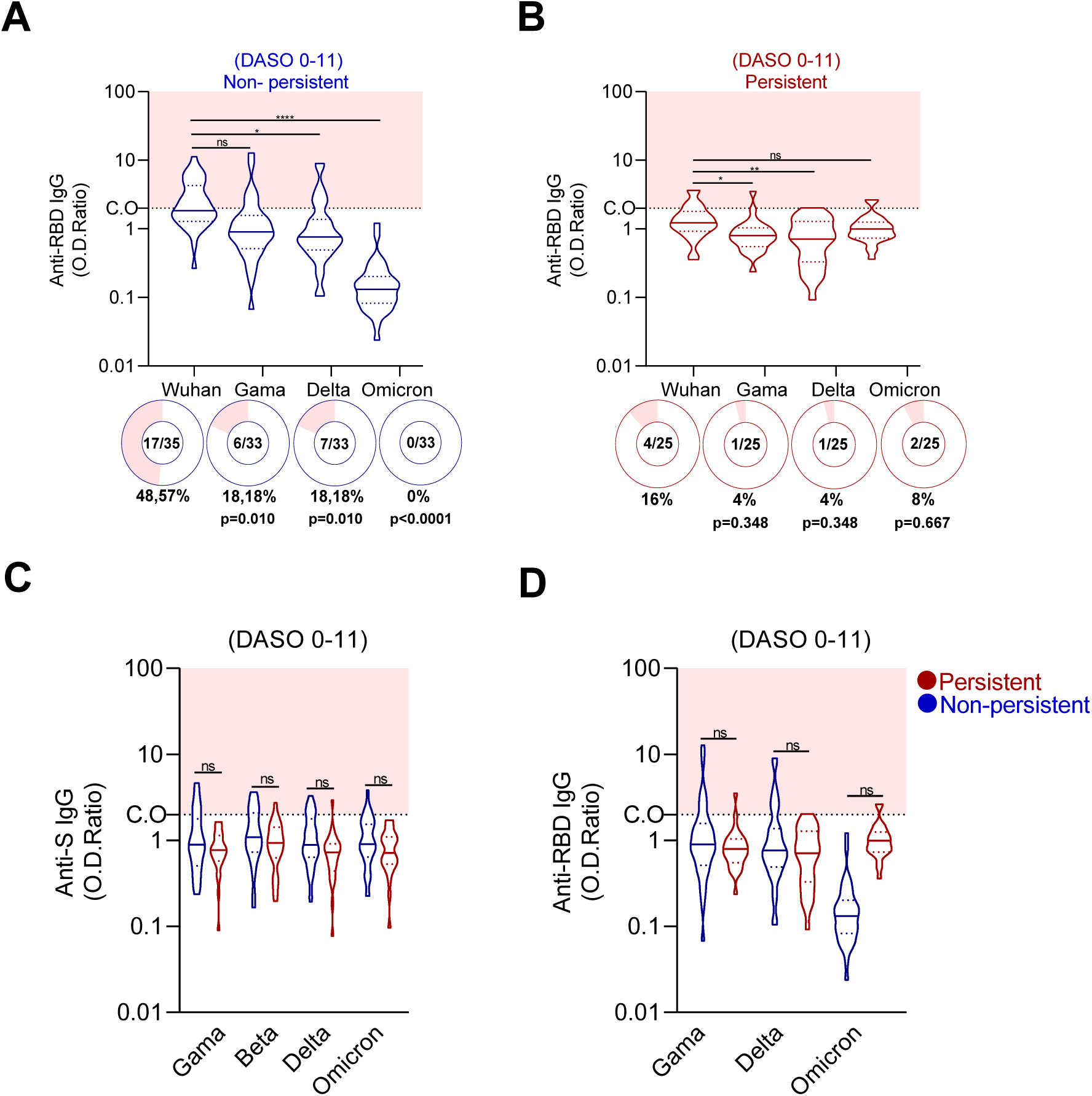
Early cross-reactive anti-Spike and anti-RBD IgG responses against SARS-CoV-2 variants in persistent and non-persistent infection. **(A–B)** Early cross-reactive anti-RBD IgG responses (0–11 DASO) against the ancestral Wuhan strain and variants of concern (Gamma, Delta, and Omicron) within the non-persistent (blue) and persistent (red) groups. The horizontal dashed line (CO) indicates the cutoff value for seropositivity, while the pink shaded area highlights the range of positive results. Donut charts show the proportion (n/N) and percentage of seropositive individuals in each group, with corresponding p-values indicating statistical comparisons of seroprevalence relative to the ancestral Wuhan strain. **(C–D)** Direct inter-group comparison of early cross-reactive anti-S and anti-RBD IgG levels between non-persistent (blue) and persistent (red) individuals across specific variants. Statistical significance: ns (not significant); * (p < 0.05); ** (p < 0.01); **** (p < 0.0001).

**Supplementary Figure 4:**
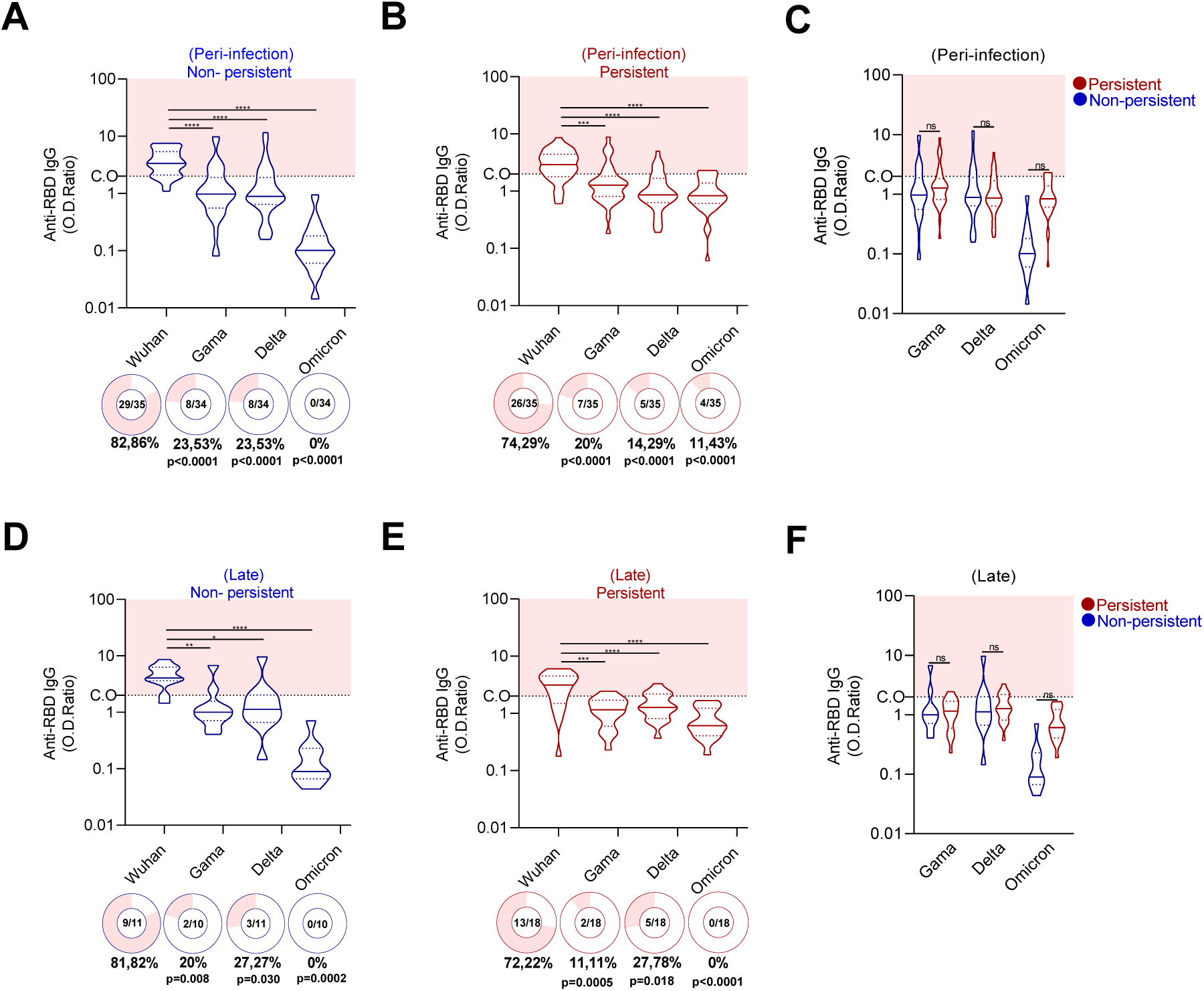
Cross-reactive anti-RBD IgG responses to SARS-CoV-2 variants during peri-infection and late convalescence. **(A–B)** Relative levels (OD ratio) of cross-reactive anti-RBD IgG against the ancestral Wuhan strain and variants (Gamma, Beta, Delta, Omicron) in non-persistent (blue) and persistent (red) individuals during the peri-infection phase. **(C)** Inter-group comparison of antibody levels during the peri-infection phase. **(D–E)** Cross-reactive anti-RBD IgG responses in both groups during the late convalescence phase, analyzed using the same criteria as in (A–B). **(F)** Inter-group comparison during the late phase. Donut charts show the proportion of seropositive individuals, with p-values for group comparisons. Statistical significance: ns; * (p < 0.05); ** (p < 0.01); *** (p < 0.001); **** (p < 0.0001)

